# A review on human gut microbial diversity in 21 tribal populations from India

**DOI:** 10.1101/2023.11.03.23297350

**Authors:** Sayak Chakraborty, Sahid Afrid Mollick, Bidyut Roy

**Affiliations:** Physical Anthropology Section, Anthropological Survey of India, Sector V, EN: 7-9, Salt Lake City, Kolkata 700091, India

**Keywords:** Gut microbiota, healthy tribal populations, India

## Abstract

From the earlier to modern times, the human populations passed through stages of subsistence such as foraging, rural farming and industrialized urban life. Till date various tribal people are living in geographically isolated areas depending on their traditional (gathering/rural farming) food sources. The long term cultural practices and food habits shaped the gut microbiome composition in these populations and contributed to health. However, the population-level study of the gut microbiota in Indian tribes with representative sampling across its geography and subsistence are limited. So, it is important to understand the complexity and diversity of the gut microbiome in healthy tribal populations, studied so far, to explore relationship among food, gut microbiome and health. Here, we reviewed gut microbiome studies which included 21 Indian tribal populations from different geographical regions to understand the role of food, ecology and cultural habits on variation of gut microbiota.

## Introduction

The human gut microbiome consists of millions of distinct bacterial species [1] which have given rise to one of the most complex ecosystems. Previous estimates of bacteria present within an individual suggested that bacterial cells outnumber human cells by the ratio 150:1, but more recent estimates put the ratio at a more moderate level of 1:1 [2]. Human microbes predominantly present in oral cavity and gut, where the majority of digestion and absorption of nutrients takes place. They consist of bacteria, along with some archaea, eukaryotes, and viruses [3] which co-evolved with their hosts to form intricate and mutually beneficial partners [4]. Over the lifetime; diversity and abundance of gut microbiome vary depending on health, environment, diet, diseases, age of individuals and other life style factors [5, 6, 7].

In India, human gut microbiota is fairly a recent domain of research which were conducted to find diversity, variation with age [8], diet [9], obesity [10], and other diseases such as irritable (inflammatory) bowel syndrome [11]. Although few studies had been conducted in Indian tribes, it was realized that India warrants vigorous and comprehensive research into the gut microbiome in diverse ethnic populations in relation to geographies, culture and customs and food and drink habits [12]. Numerous pre-industrial and pre-agricultural tribal societies exist in India which has been earmarked by the Government of India. Among total 705 Scheduled Tribes (ST); 75 are identified as Particularly Vulnerable Tribal groups (PVTGs) [13]. These PVTGs, to an extent, have maintained much of their traditional lifestyle and social practices, so, investigations of gut microbiome in healthy PVTGs would be very useful data to understand health status in them. In this review, we will enumerate gut microbial diversity in healthy Indian tribes, studied so far.

## Materials and Methods

Previously published studies related to gut microbiome of healthy Indian tribal communities were searched. The entire process of exclusion and inclusion process of the research papers reviewed in this study have been shown in a flowchart (Figure 1) prepared using PRISMA 2020 template. Different headings and keywords were used to search the articles; such as ‘tribe’, ‘India’, ‘human gut microbiota’, ‘human gut microbiome’, ‘metagenomics’, and ‘human gut microbial diversity’. A total number of 36 published articles were identified from electronic databases such as Research Gate (n=1), and metadata services such as PubMed (n=25) and Google Scholar (n=10). From these 36 articles, 14 records were removed before screening since they did not match the keywords perfectly, so a total of 22 papers were selected for scrutiny. Again, one article was excluded due to the lack of proper description of quantitative data and access to its supplementary files, so, 21 papers (8 from PubMed, 1 from Research Gate and 12 from Google Scholar) were considered for the scrutiny. Further, a total of 16 papers were excluded for the reasons such as 4 articles overlapped with separate online sources, 8 articles had insufficiency for the objective of the present review, and 4 articles did not have clear names of the tribal populations. Finally, 5 articles fulfilled our inclusion criteria and, hence, considered for the review and these publications reported 22 tribal populations and 1 non-tribal rural population across 7 states of India (Figure 2; Table 1). But data from this non-tribal population was not considered in this review.

**Figure 1.**
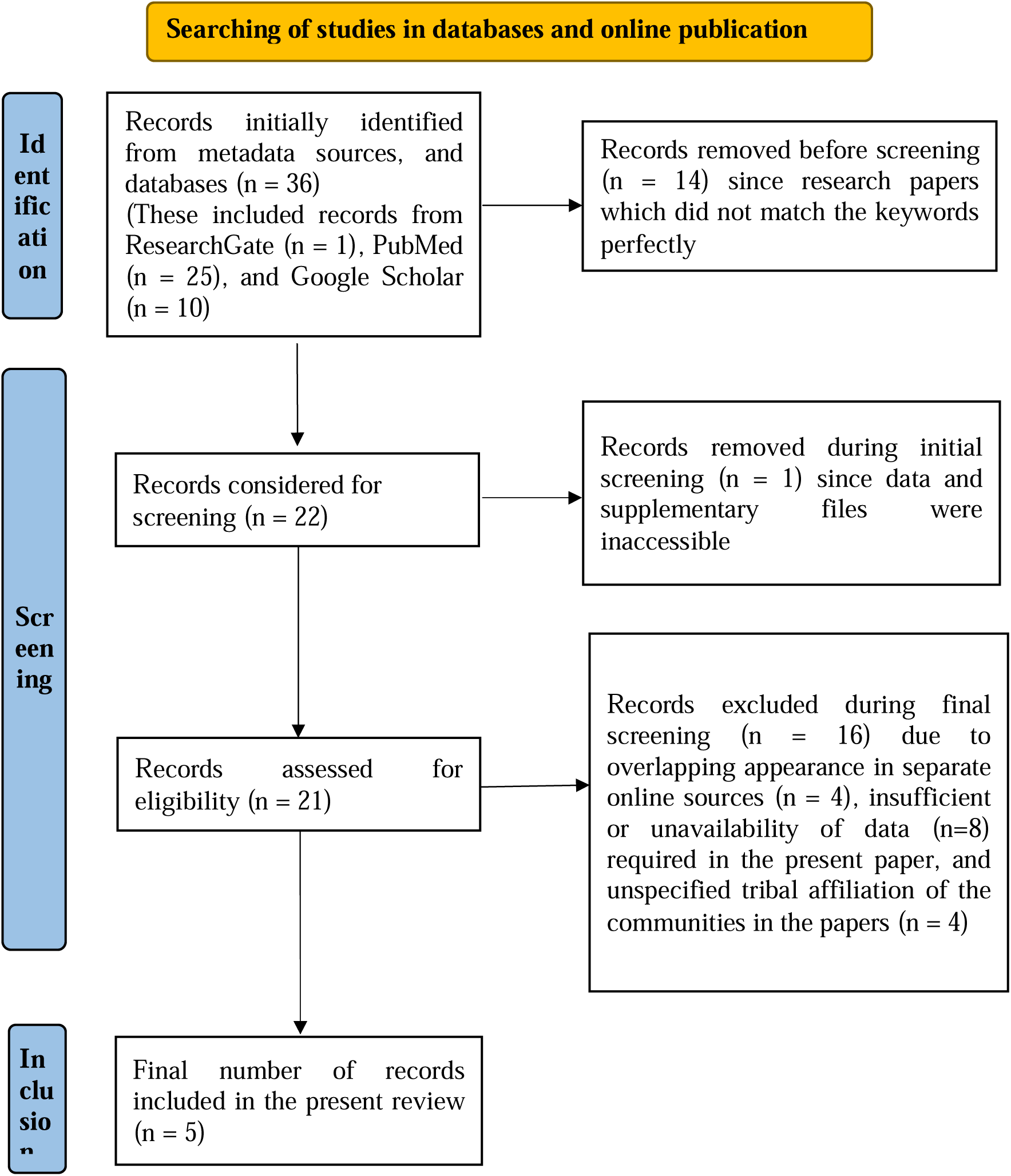
Flowchart for the selection of the research papers considered in the present review.

**Figure 2.**
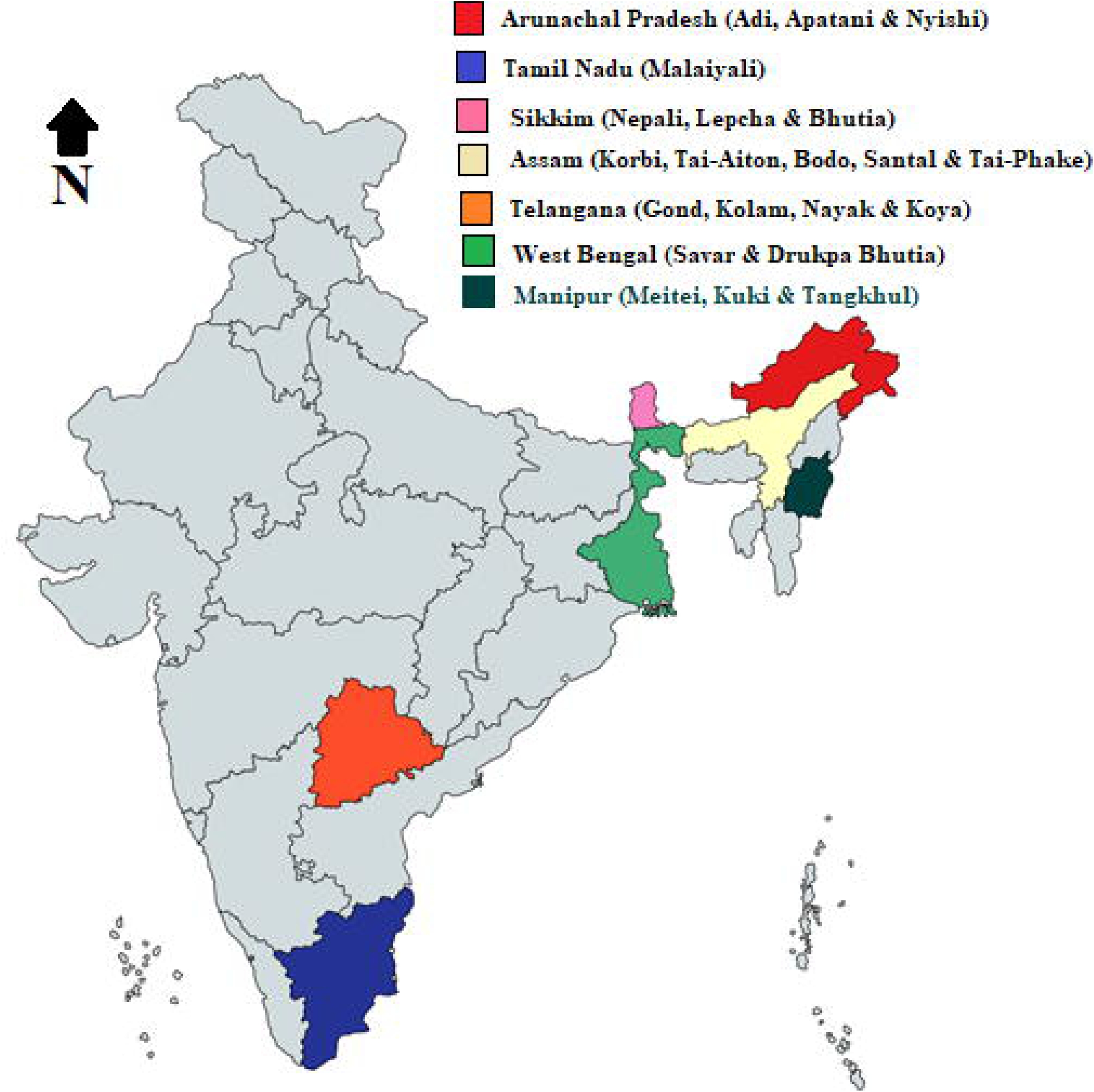
Geographical locations/states of the tribal communities whose gut microbiome has been studied.

**Table 1.** Specifics of the studies reviewed here.

| Sl. No. | Publications used in this review | Studied Tribal Communities (21+1) | Sample Size | Region of the Study | Diet/Lifestyle/Mode of Subsistence | Differential Composition of bacteria (Phylum-level) | Proposed Influential Factors | Remarks, or Outcomes of the Study |
| --- | --- | --- | --- | --- | --- | --- | --- | --- |
| 1. | Dehingia et al. 2015 [14] | Korbi, Tai-Aiton, Bodo, Santal, Tai-Phake | 78 | Assam | Rice is the staple food for all the tribal groups, with variations in consumption of vegetables, fish, meat, legumes, whole grains, fruits, and tubers. The tribes from Manipur and | *Firmicutes*,<br>*Bacillota*,<br>*Bacteroidetes*,<br>*Actinomycetota*,<br>*Proteobacteria* | Geography and diet | Geography and diet had significant effect on the gut bacterial profile of these Indian tribes which were dominated by *Prevotella* |
|  |  | Meitei, Kuki, Tangkhul | 30 | Manipur |  |  |  |  |
|  |  | Nepali, Bhutia, Lepcha | 27 | Sikkim |  |  |  |  |
|  |  | Gond, Kolam,<br>Nayak, Koya | 58 | Telangana, | Sikkim consume a relatively higher quantity of fermented foods, and dairy, along with dried and smoked fish and meat. |  |  |  |
|  |  | Total | 193 |  |  |  |  |  |
| 2. | Ramadass et al. 2017 [15] | One Rural Population (Unknown Ethnic Group) | 10 | Tamil Nadu | Consume millets-based diet. Eat pork every day. Do not consume any dairy products. | *Firmicutes, Proteobacteria, Bacteroidetes, Actinobacteria* | Dietary habits | *Actinobacteria* were significantly low in this group. |
|  |  | One Rural Tribal Population (Malaiyali) | 10 |  |  |  |  |  |
|  |  | Total | 20 |  |  |  |  |  |
| 3. | Ganguli et al. 2019 [16] | Savar | 3 (father, mother, and child) | West Bengal | Foragers, (essentially 'carnivores') and dependent on forest produce. | *Verrucomicrobiota, Bacillota, Actinomycetota, Bacteroidetes, Firmicutes* | Geographic location and dietary habits | The propensity of transition of the gut microbial profiles from parents to son. Thus, revealing the parental contribution to the formation of the child's gut microbiome. |
| 4. | Basu et al. 2022 [17] | Drukpa Bhutia | 3 (father, mother, and child) | West Bengal | Intake of locally produced vegetables and fruits, along with liquor and fermented dairy products. | *Bacteroidetes, Actinomycetota, Bacillota, Firmicutes, Pseudomonadota* | Traditional dietary practice | Parental contribution in the formation of the child's gut microbiome. |
| 5. | Hazarika et al. 2022 [18] | Adi | 10 | Arunachal Pradesh | The Nyshi consume cereals, millet, leaves, fish, and meat. The Apatani tribe consume boiled vegetables, boiled fish, meat, and dairy products. All three tribes also consume fermented bamboo shoots, smoked pork, and smoked fish. Rice is their main staple cereal. | *Verrucomicrobiota*,<br>*Firmicutes*,<br>*Actinobacteria*,<br>*Proteobacteria*,<br>*Fusobacteria*,<br>*Elusimicrobia*,<br>*Bacteroidetes* | Dietary factors, and the overall state of health | Higher gut microbiome diversity with a high prevalence of *Prevotella* and *Collinsella* in the Adi and the Nyshi. *Bifidobacterium* and *Catenibacterium* in the Apatani. |
|  |  | Apatani | 10 |  |  |  |  |  |
|  |  | Nyshi | 10 |  |  |  |  |  |
|  |  | Total | 30 |  |  |  |  |  |

In these studies, fecal samples were collected aseptically from all individuals, kept at -20° or -80 ° as reported and used for bacterial DNA isolation using different methods (Table 2). Segment of bacterial ribosomal 16S DNA were PCR amplified and sequenced in different NGS platforms to find the diversity of gut bacteria in individuals of these populations. Analyzed data of the phyla and genera of the bacteria were taken from the reported papers. Heat maps of the bacterial phyla and genera of the samples were tabulated in MS Excel 2010 (Figure 3, Figure 4) using PAST 4.04.

**Figure 3.**
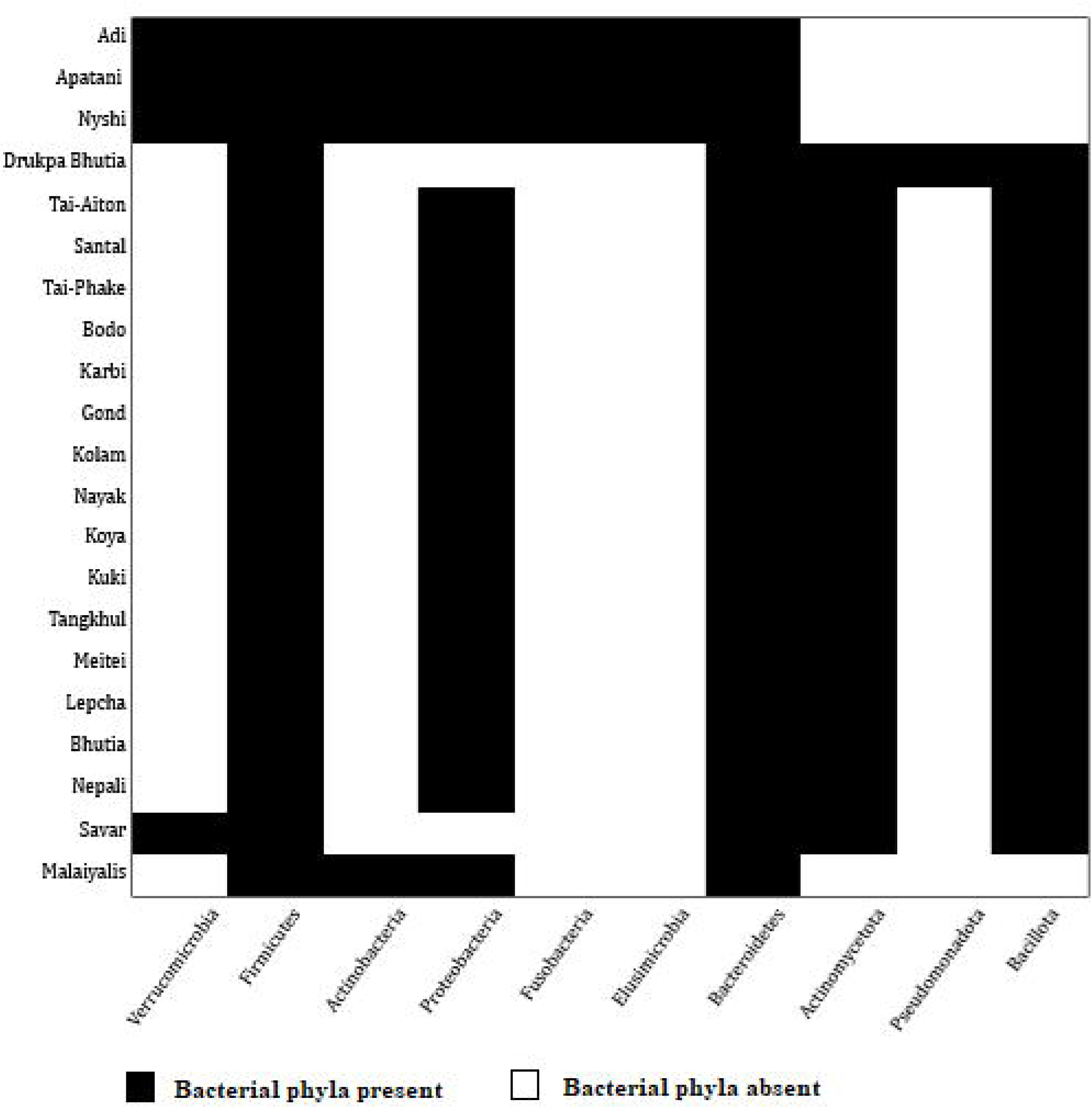
Phylum of the bacteria present in the studied tribes.

**Figure 4.**
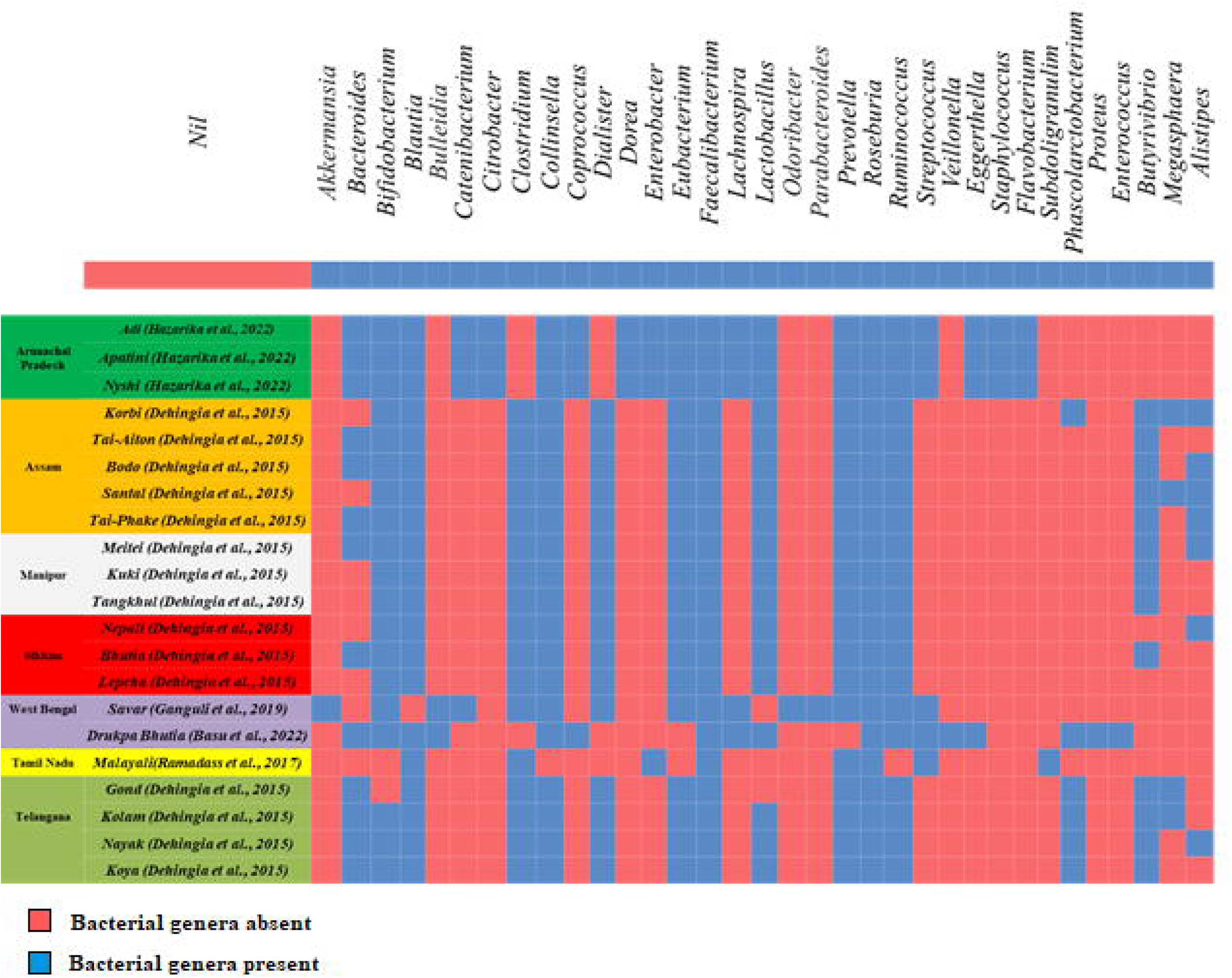
Genus of the bacteria present in the studied tribes.

**Table 2.** Methods employed in the reported studies.

| Sl. No. | Publications | Sample size (Age ranges) | Study Objectives | Specimen Type | Techniques |
| --- | --- | --- | --- | --- | --- |
| 1. | Dehingia et al. 2015 [14] | 193 (20-35 years) | Gut bacterial diversity of the tribes | Faecal samples | DNA was extracted from the faecal samples using QIAGEN DNA Stool Mini-Kit (QIAGEN, Hilden, Germany). Sequencing of the V3- V4 regions of 16S rRNA was done on the Illumina MiSeq Platform. |
| 2. | Ramadass et al. 2017 [15] | 20 (18-60 years) | Comparison of faecal microbiota of healthy adults in south India among the tribal and non-tribal | Faecal samples | DNA was extracted from approximately faecal samples using QIAamp DNA stool mini kits. Sequencing the V3-V4 regions of 16s rRNA was done using Roche 454 GLX Titanium |

|  |  |  | population |  | sequencer (Roche Diagnostics GmbH) |
| --- | --- | --- | --- | --- | --- |
| 3. | Ganguli et al. 2019<br>[16] | 3 (7, 26, and<br>30 years) | Gut microbiome of a foraging<br>tribe. | Faecal<br>samples | DNA isolation was done following the THSTI<br>Method given by Bag et al. (2016) [19]. NGS<br>Sequencing was done in the Illumina Hiseq<br>sequencing platform. |
| 4. | Basu et al. 2022<br>[17] | 3 (5, 27, and<br>29 years) | Gut microbiome of a tribal<br>population | Faecal<br>samples | DNA isolation was done following the THSTI<br>Method given by Bag et al. (2016) [19]. NGS<br>Sequencing was done in the Illumina Hiseq<br>sequencing platform. |
| 5. | Hazarika et al.<br>2022 [18] | 30 (20-60<br>years) | Gut microbiome of a tribal<br>community | Faecal<br>samples | DNA was extracted from stool samples using<br>the QIAamp DNA Stool Mini Kit. Sequencing<br>of the V3- V4 regions of 16S rRNA was done<br>on the Illumina MiSeq sequencing platform. |

## Analysis and Results

All reported articles mainly attempted to address the gut microbial diversity in these tribal populations based on three major factors such as dietary and cultural habits, ethnicity, and geographic location (Table 1). Diet and social drinks has been found to be different in different tribes based on their geography and ethnicity. The North-East Indian tribes, including Sikkim, were found to have similar food habits, though they belongs to different geo-political boundaries and shared ethnic affiliation. Sample sizes in the studies were different ranging from 193 [14] to merely 3 [16, 17].

Thirty-four bacterial genera, belonging to 10 phyla, have been reported in these studies (Figure 3 and Figure 4). Except Hazarika et al. (2022) [18], remaining studies reported presence of 3 bacterial phyla *Bacteroidetes* (generally known by its more popular synonym *Bacteroidota)*, *Firmicutes*, and *Bacillota*. However, *Bacillota* is presently considered to be a synonym of *Firmicutes* [20]. So, only 2 phyla (*Bacteroidota* and *Bacillota)* were found across all the studied populations. Apart from the above-mentioned 2 phyla, 8 other bacterial phyla (*Actinomycetota, Proteobacteria*, *Actinobacteria*, *Spirochaetes, Elusimicrobia, Pseudomonadota, Verrucomicrobiota,* and *Fusobacteria)* have been reported in these tribal populations, irrespective of ethnicity, geography and dietary habits. *Firmicutes, Proteobacteria,* and *Bacteroidetes* were the most abundant bacterial phyla reported in these populations. Additionally, few North-East Indian tribes (Adi, Apatani, Nyshi) and one South Indian tribal group (Malaiyali) had *Actinobacteria.* One tribe (Drukpa Bhutia) from West Bengal had *Pseudomonadota*. The phylum *Fusobacteria* were only found in North-East Indian tribes (Adi, Apatani, and Nyshi) from Arunachal Pradesh (Figure 3).

At the genus-level, the most abundant bacteria were *Faecalibacterium* and *Roseburia* (both from phylum *Bacillota*) which have been reported in all the tribal populations mentioned here. The bacterial genera of *Ruminococcus* (phylum *Bacillota*)*, Prevotella* (phylum *Bacteroidetes*)*, Blautia* (phylum *Bacillota*) were present in nearly all studied populations, followed by *Bifidobacterium* (phylum *Actinomycetota*) and *Eubacterium* (phylum Bacillota). The bacterial genera with minimal presence, (i.e. occurring in only one tribal community) were: *Akkermansia* (in Savar), *Odoribacter* (in Savar), *Parabacteroides* (in Savar)*, Veillonella* (in Drukpa Bhutia), *Subduligranulim* (in Malaiyali), *Proteus* (in Drukpa Bhutia) and *Enterococcus* (in Drukpa Bhutia) (Figure 4).

The core gut bacteria in the tribes of Telangana and Assam consisted of *Prevotella*, *Faecalibacterium*, *Eubacterium*, *Clostridium*, *Blautia*, *Collinsella*, *Ruminococcus* and *Roseburia*. Additionally, the bacterial genera *Bacteroides*, *Dialister* and *Veillonella* were also found to be core bacteria in the tribes of Manipur. *Bacteroides*, *Dialister*, *Bifidobacterium* and *Lactobacillus* were the genera found in the tribes of Sikkim. The Adi and Nyshi tribes of Arunachal Pradesh had high prevalence of *Prevotella* and *Collinsella*, but Apatani tribe had high prevalence of *Bifidobacterium* and *Catenibacterium*, even though these tribes were living in the same geographical region and had similar dietary practices. In Savar and Drukpa Bhutia tribes from West Bengal; *Bacteroides*, *Bifidobacterium*, *Streptococcus*, *Faecalibactenum*, *Ruminococcus*, *Prevotella*, *Roseburia*, *Collinsella* were the most common bacterial genus found in both the communities despite they were living in separate geographic regions of the state and had different dietary practices (Figure 2 and Figure 4).

## Discussion

India has become a melting pot where the different lifestyle factors and food choices are being infiltrated in many indigenous and traditional lifestyles even in tribal communities [14] with the alteration of the gut microbiome profile [16, 18]. Although some communities still remained isolated so there are some chances to study gut bacterial diversity in these isolated tribal populations to compare the future changes in gut bacterial diversity due to food and life styles factors. Striking disparities between the gut microbial taxa of urban populations and rural tribal populations have also been reported [21] which indicate such shifts may have been caused by different dietary and lifestyle practices. Dehingia et al. (2015) [14] claimed that geographical locations, culture and diet had significant influence on gut bacterial profile of the Indian tribes which were dominated by the genus *Prevotella*. The Drukpa Bhutia tribe in north of West Bengal followed traditional agriculture based diet along with some foraging and livestock rearing products [17] and the Savar tribe in south of West Bengal led a relatively more foraging lifestyle [16], but both of them retained their ancestral gut microbiome profile since they were not impacted much by external influences such as present-day life style factors and food. These two studies, on Drukpa Bhutia and Savar, also claimed parental contribution to the composition of the child’s gut microbiota.

The Adi, Apatani, and Nyshi communities seemed to have same bacterial phyla (Supplementary Table 1, Figure 3) and genera (Supplementary Table 1, Figure 4) in their gut. This might be due to similar dietary patterns supported by agricultural subsistence economy (Table 2) and shared highland ecology of Arunachal Pradesh. In addition, all of them share a common ancestral cultural identity given that they are all speakers of the Tani sub-group of languages in the Sino-Tibetan language family [22]. These three indigenous groups from Arunachal Pradesh are also unique in the context that they had the highest diversity of phyla among all indigenous studied tribes (Figure 3). An interesting observation was that apart from the three aforementioned tribes of Arunachal Pradesh; the Drukpa Bhutia and the Savar from West Bengal; the Malaiyalis fromTamil Nadu and other indigenous communities, spread across the states of Manipur, Assam, Telangana, and Tamil Nadu contained the same five bacterial phyla (*Firmicutes*, *Proteobacteria*, *Bacteroidetes*, *Actinomyctetota*, and *Bacillota)* (Figure 3) in their gut samples. So, similarities of phyla were observed in tribal populations living over such a large ecological range and unrelated linguistic communities (such as Tibeto-Burmese, Dravidian, and Austro-Asiatic groups) but there were differences at the level of the bacterial genera in these populations (Figure 4).

*Faecalibacterium* and *Roseburia* (both from phylum *Bacillota*) have been reported to be present in the gut of all the tribal groups. *Faecalibacterium* (its sole species, *F. prausnutzii*) is known to be a beneficial bacterium in the human gut as it produces butyrate (a small chain fatty acid) in the intestine for the metabolism of sugar. Further, it helps in protecting diseases of the bowel such as Crohn’s Disease [23] and irritable bowel disease [24]. *Roseburia* has similar functions like *Faecalibacterium*. For optimum performance, *Faecalibacterium* requires presence of some other bacteria such as *Bacteroides thetaiotaomicron* in gut [25] in order to make more butyrate production by *Faecalibacterium.* Among the populations mentioned in the review, 14 out of 21 tribes had *Bacteroides* alongside *Faecalibacterium* in gut.

The tribes in Telangana [14] seem to have the same 5 bacterial phyla (*Firmicutes*, *Proteobacteria*, *Bacteroidetes*, *Actinomycetota*, and *Bacillota*) in their guts and share 18 similar bacterial genera. Among them; Gond, Kolam, and Koya are Dravidian-speaking communities which are enlisted as Scheduled Tribes of India, however the identity of the fourth tribe, Nayak, is not clear. Nayak or Naik in Telangana is a surname adopted by various communities [26] and may not be a single tribal identity. The gut microbial composition of three Tibeto-Burmese tribes (i.e. Nepali, Bhutia, and Lepcha) of Sikkim showed presence of 5 common bacterial phyla (*Firmicutes*, *Proteobacteria*, *Bacteroidetes*, *Actinomycetota*, and *Bacillota*) (Figure 4) and 20 common genera. It may be noted that bacterial phyla present in the tribes of Telangana and Sikkim are similar, despite their geographic, culture, dietary and ethnic differences.

The Savars of West Bengal happened to have three genera of bacteria unique to them: *Akkermansia* (phylum *Verrucomicrobia*), *Odoribacter* and *Parabacteroides* (both from phylum *Bacteroidetes*). *Akkermansia* has been reported to have several benefits towards human health such as reduction of body weight, resistance towards bowel diseases, and even cancer [27]. A decreased level of *Odoribacter* in the human gut has been related to various microbiota-associated diseases such as non-alcoholic fatty liver, cystic fibrosis, and inflammatory bowel disease [28]. Abundance of *Parabacteroides* in the gut is known to be useful in the fight against obesity and other gut-associated diseases [29, 30, 31]. On the other hand, the Drukpa Bhutia from the northern part of West Bengal had unique genera: *Veillonella*, *Enterococcus* (phylum *Bacillota*), and *Proteus* (phylum *Pseudomonadota*). But *Veillonella* helps in enhancement of performance of individuals via lactic acid metabolism and production of sugar [32]. The Drukpa Bhutia inhabits at remote regions of semi-forested land. The occurrence of a bacterium associated with work performance enhancement may be related to the dairy consumption which is a key source of sugar for energy. *Enterococcus* is infamous for its virulence and association with various diseases such as sepsis, urinary tract infections, and meningitis within the human body [33]. Its occurrence in the gut of Drukpa Bhutia did not reveal any associated disease and may require further detailed investigation with more samples and involvement of clinicians. Similarly, *Proteus* is another pathogenic bacterium and causes urinary tract infections, so further investigation is very urgent.

While considering the details of the research papers reviewed, it was noticed that these studies had several limitations. Ramadass et al. (2017) [15] used two broad sample groups, one consisting of individuals from the Malaiyali community (termed as a ‘rural tribal population’) and a ‘rural population’. The “rural population” hints a generic term used to designate individuals of a non-homogeneous non-tribal group consisted of individuals from different religion, caste, etc. Using such a sample of non-homogeneous rural inhabitants to compare with one single homogeneous tribal community, inhabiting the same region/district in Tamil Nadu, may yield the spurious results. So, data of this “rural population” was excluded from this review. After all; abundance, composition, richness and evenness of human gut microbiota depends on lifestyle and dietary pattern specific to communities. Dehingia et al. (2015) [14] used a large sample size (n = 193) from 15 tribal communities. But they mentioned that these tribal communities belonged to different states with low sample size in each state: Assam (5 communities, n = 78), Manipur (3 communities, n = 30), Sikkim (3 communities, n = 30), and Telangana (4 communities, n = 58). The lack of availability of sample size of each community and non-uniformity of sample size in each community limited the study for comparing tribal specific gut microbiome. Those studies seem to be preliminary reports since none of them mentioned the sample size required for this type of study with sufficient power. In case of the studies on the Savar [16] and Drukpa Bhutia [17], from West Bengal, the sample sizes were very low (n = 3) and constituted a single nuclear family unit (mother, father and a child). These two papers aimed at visualizing the diversity of bacterial phyla and genera in the guts of these families and discussing the passage of the bacterial genera/taxa from parents to child in the same family. The researchers should have considered several nuclear family units of mother, father, and child to get proper statistical power and similarity index (Czekanowski Coffiecient and Squared Euclidean Distance) to know the passing of bacterial taxa and genera from parents to child.

## Conclusion

Healthy tribal groups of India are important populations to find bacterial diversity in gut microbiota since most of them living in isolated places without much mixing with urban populations. Relative similarity in the core gut microbiota of tribes was noted, considering their close geographical proximity, similar dietary practices and common ethno-linguistic group. Although, Chaudhari et al. (2020) [34] reported association of age with the frequencies of different gut bacteria, but we could not study changes of bacterial genus as the age changes since in these populations since sample sizes were low and detailed demographic details were not available. Further studies on gut microbiota of isolated tribal groups can help to reveal the role of gut microbiota towards the health and predisposition towards gastrointestinal diseases and discovery of probiotic markers for therapeutic purpose.

## Author Contribution

SC: Wrote draft of paper, Figures and Tables, SAM: Initial draft, searching of related review articles, Initial Figures and Tables; BR: Final paper, Figures and Tables

## Data Availability

all data shown in the present work are contained in the manuscript

## Acknowledgement

SC and SAM received SRF from Anthropological Survey of India during this study. BR: Received R N Tagore Scholarship from Ministry of Culture, Government of India to work at Anthropological Survey of India, Kolkata.

---

**Supplementary Table 1:**
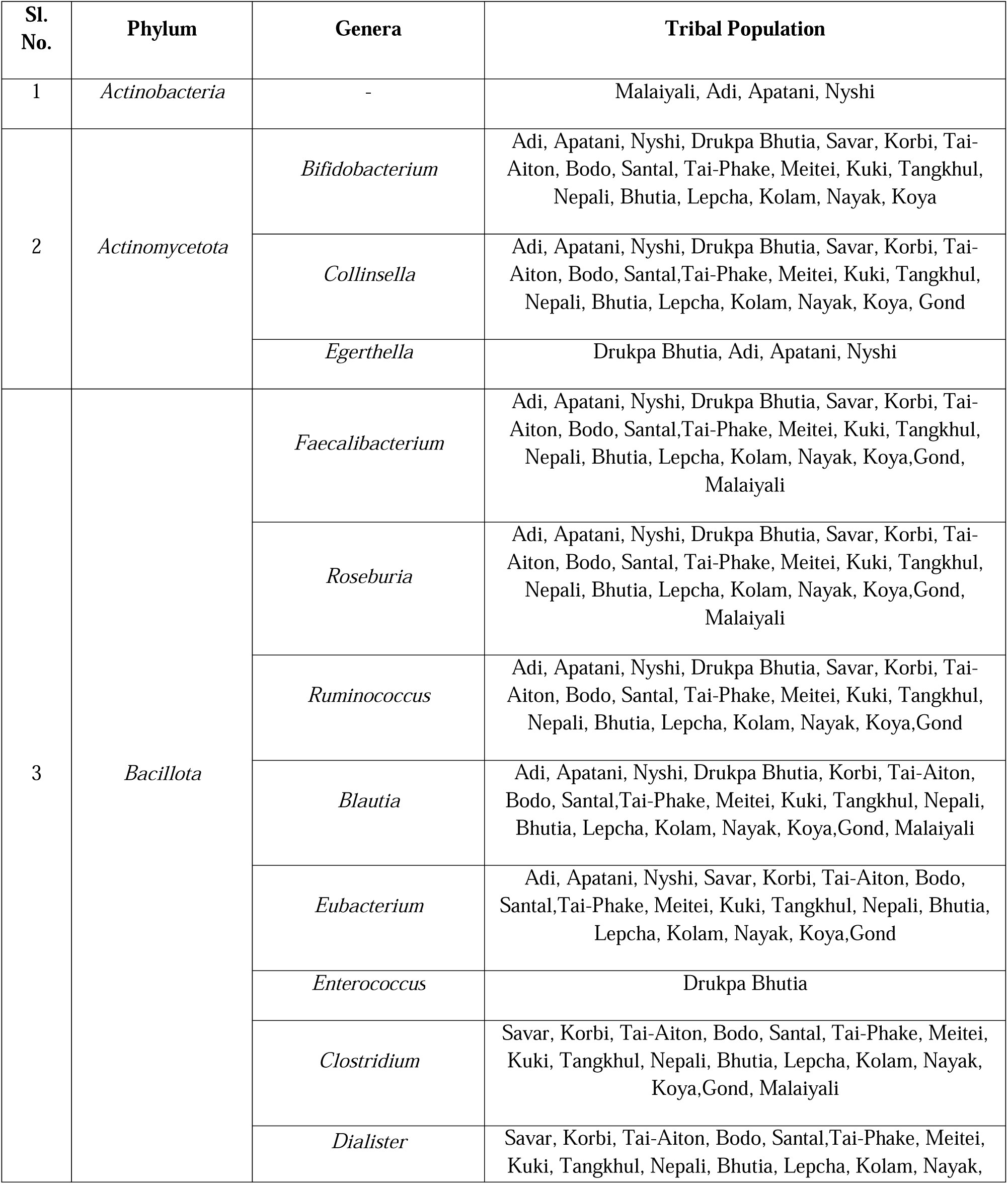
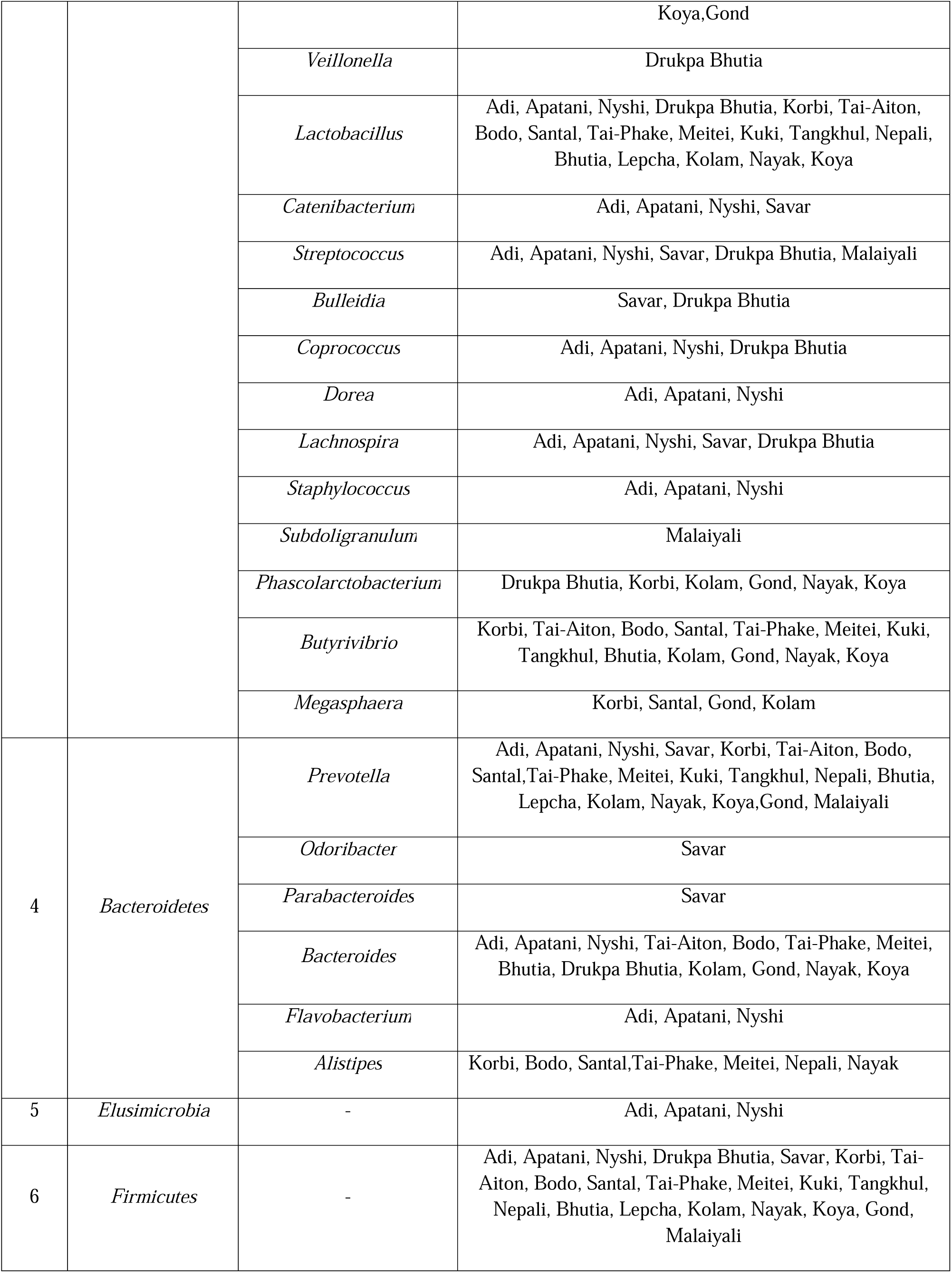
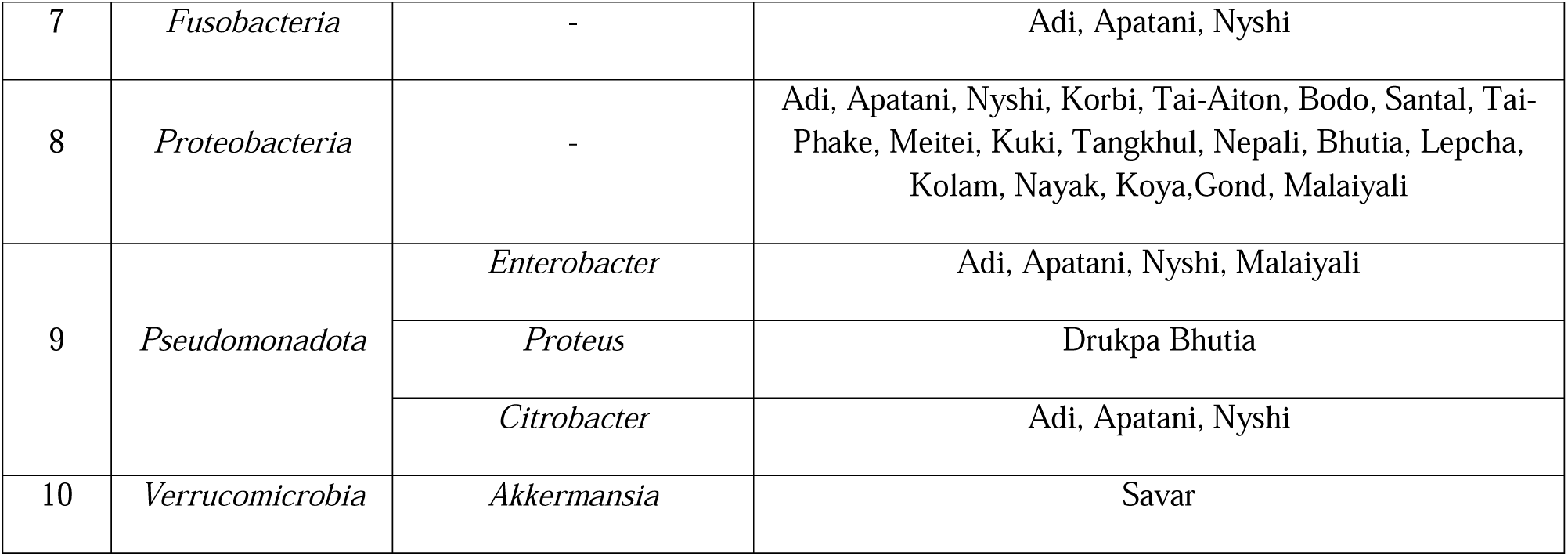
Phylum and genera of the various gut microbes present in the studied tribal populations.

## References

1. Sankar, S.A., Lagier, J.C., Pontarotti, P., Raoult, D. and Fournier, P.E., 2015. The human gut microbiome, a taxonomic conundrum. Systematic and Applied Microbiology, 38(4), p.276–286.

2. Sender, R., Fuchs, S. and Milo, R., 2016. Are we really vastly outnumbered? Revisiting the ratio of bacterial to host cells in humans. Cell, 164(3), p.337–340.

3. Matijašić, M., Meštrović, T., Čipčić Paljetak, H., Perić, M., Barešić, A. and Verbanac, D., 2020. Gut microbiota beyond bacteria—mycobiome, virome, archaeome, and eukaryotic parasites in IBD. International Journal of Molecular Sciences, 21(8), p.2668.

4. Thursby, E. and Juge, N., 2017. Introduction to the human gut microbiota. Biochemical Journal, 474(11), p.1823–1836.

5. Yatsunenko, T., Rey, F.E., Manary, M.J., Trehan, I., Dominguez-Bello, M.G., Contreras, M., Magris, M., Hidalgo, G., Baldassano, R.N., Anokhin, A.P. and Heath, A.C., 2012. Human gut microbiome viewed across age and geography. Nature, 486(7402), p.222–227.

6. Karlsson, F., Tremaroli, V., Nielsen, J. and Bäckhed, F., 2013. Assessing the human gut microbiota in metabolic diseases. Diabetes, 62(10), p.3341–3349.

7. David, L.A., Maurice, C.F., Carmody, R.N., Gootenberg, D.B., Button, J.E., Wolfe, B.E., Ling, A.V., Devlin, A.S., Varma, Y., Fischbach, M.A. and Biddinger, S.B., 2014. Diet rapidly and reproducibly alters the human gut microbiome. Nature, 505(7484), p.559–563.

8. Marathe, N., Shetty, S., Lanjekar, V., Ranade, D. and Shouche, Y., 2012. Changes in human gut flora with age: an Indian familial study. BMC Microbiology, 12, p.1–10.

9. Kabeerdoss, J., Jayakanthan, P., Pugazhendhi, S. and Ramakrishna, B.S., 2015. Alterations of mucosal microbiota in the colon of patients with inflammatory bowel disease revealed by real time polymerase chain reaction amplification of 16S ribosomal ribonucleic acid. The Indian Journal of Medical Research, 142(1), p.23.

10. Patil, D.P., Dhotre, D.P., Chavan, S.G., Sultan, A., Jain, D.S., Lanjekar, V.B., Gangawani, J., Shah, P.S., Todkar, J.S., Shah, S. and Ranade, D.R., 2012. Molecular analysis of gut microbiota in obesity among Indian individuals. Journal of Biosciences, 37, p.647–657.

11. Verma, R., Verma, A.K., Ahuja, V. and Paul, J., 2010. Real-time analysis of mucosal flora in patients with inflammatory bowel disease in India. Journal of Clinical Microbiology, 48(11), p.4279–4282.

12. Shetty, S.A., Marathe, N.P. and Shouche, Y.S., 2013. Opportunities and challenges for gut microbiome studies in the Indian population. Microbiome, 1(1), p.1–12.

13. Ministry of Home Affairs, Government of India. 2001, 2011. Census of India.

14. Dehingia, M., Thangjam Devi, K., Talukdar, N.C., Talukdar, R., Reddy, N., Mande, S.S., Deka, M. and Khan, M.R., 2015. Gut bacterial diversity of the tribes of India and comparison with the worldwide data. Scientific Reports, 5(1), p.18563.

15. Ramadass, B., Rani, B.S., Pugazhendhi, S., John, K.R. and Ramakrishna, B.S., 2017. Faecal microbiota of healthy adults in south India: comparison of a tribal & a rural population. The Indian Journal of Medical Research, 145(2), p.237.

16. Ganguli, S., Pal, S., Das, K., Banerjee, R. and Bagchi, S.S., 2019. Gut microbial dataset of a foraging tribe from rural West Bengal-insights into unadulterated and transitional microbial abundance. Data in Brief, 25, p.103963.

17. Basu, S., Das, K., Ghosh, M.M., Banerjee, R., Bagchi, S.S. and Ganguli, S., 2022. First report of gut bacterial dataset of a tribal Bhutia family from West Bengal, India. Data in Brief, 41, p.107859.

18. Hazarika, P., Chattopadhyay, I., Umpo, M., Choudhury, Y. and Sharma, I., 2022. Elucidating the gut microbiome alterations of tribal community of Arunachal Pradesh: perspectives on their lifestyle or food habits. Scientific Reports, 12(1), p.18296.

19. Bag, S., Saha, B., Mehta, O., Anbumani, D., Kumar, N., Dayal, M., Pant, A., Kumar, P., Saxena, S., et al. 2016. An Improved Method for High Quality Metagenomics DNA Extraction from Human and Environmental Samples. Scientific Reports, 6, p.26775.

20. Oren, A. and Garrity, G.M., 2021. Valid publication of new names and new combinations effectively published outside the IJSEM. International Journal of Systematic and Evolutionary Microbiology, 71(11), p.005096.

21. Singh, R., Haque, M.M. and Mande, S.S., 2019. Lifestyle-induced microbial gradients: an Indian perspective. Frontiers in Microbiology, 10, p.2874.

22. Sun, T. J. (1993). A Historical-Comparative Study of the Tani (Mirish) Branch in Tibeto-Burman. University of California, Berkeley.

23. Wright, E.K., Kamm, M.A., Teo, S.M., Inouye, M., Wagner, J. and Kirkwood, C.D., 2015. Recent advances in characterizing the gastrointestinal microbiome in Crohn’s disease: a systematic review. Inflammatory Bowel Diseases, 21(6), p.1219–1228.

24. Lopez-Siles, M., Duncan, S.H., Garcia-Gil, L.J. and Martinez-Medina, M., 2017. Faecalibacterium prausnitzii: from microbiology to diagnostics and prognostics. The ISME Journal, 11(4), p.841–852.

25. Wrzosek, L., Miquel, S., Noordine, M.L., Bouet, S., Chevalier-Curt, M.J., Robert, V., Philippe, C., Bridonneau, C., Cherbuy, C., Robbe-Masselot, C. and Langella, P., 2013. *Bacteroides thetaiotaomicron* and *Faecalibacterium prausnitzii* influence the production of mucus glycans and the development of goblet cells in the colonic epithelium of a gnotobiotic model rodent. BMC Biology, 11, p.1–13.

26. Singh, K. S., 1993. Ethnography, Customary Law, and Change. Concept Publishing Company. p.249.

27. Jayachandran, M., Chung, S.S.M. and Xu, B., 2020. A critical review of the relationship between dietary components, the gut microbe Akkermansia muciniphila, and human health. Critical Reviews in Food Science and Nutrition, 60(13), p.2265–2276.

28. Hiippala, K., Barreto, G., Burrello, C., Diaz-Basabe, A., Suutarinen, M., Kainulainen, V., Bowers, J.R., Lemmer, D., Engelthaler, D.M., Eklund, K.K. and Facciotti, F., 2020. Novel Odoribacter splanchnicus strain and its outer membrane vesicles exert immunoregulatory effects in vitro. Frontiers in Microbiology, 11, p.575455.

29. Wang, K., Liao, M., Zhou, N., Bao, L., Ma, K., Zheng, Z., Wang, Y., Liu, C., Wang, W., Wang, J., Liu, S. J., Liu, H., 2019. *Parabacteroides distasonis* Alleviates Obesity and Metabolic Dysfunctions via Production of Succinate and Secondary Bile Acids. Cell Reports, 26(1), p.222–235.

30. Wu, T.R., Lin, C.S., Chang, C.J., Lin, T.L., Martel, J., Ko, Y.F., Ojcius, D.M., Lu, C.C., Young, J.D. and Lai, H.C., 2019. Gut commensal *Parabacteroides goldsteinii* plays a predominant role in the anti-obesity effects of polysaccharides isolated from *Hirsutella sinensis*. Gut, 68(2), p.248–262.

31. Zeng, Q., Li, D., He, Y., Li, Y., Yang, Z., Zhao, X., Liu, Y., Wang, Y., Sun, J., Feng, X. et al., 2019.Discrepant gut microbiota markers for the classification of obesity-related metabolic abnormalities. Sci Rep 9, 13424.

32. Scheiman, J., Luber, J.M., Chavkin, T.A., MacDonald, T., Tung, A., Pham, L.D., Wibowo, M.C., Wurth, R.C., Punthambaker, S., Tierney, B.T. and Yang, Z., 2019. Meta-omics analysis of elite athletes identifies a performance-enhancing microbe that functions via lactate metabolism. Nature Medicine, 25(7), p.1104–1109.

33. Murray, B E., 1990. The life and times of the Enterococcus. Clinical Microbiology Reviews, 3(1), p.46–65.

34. Chaudhari, D. S., Dhotre, D. P., Agarwal, D. M., Gaike, A. H., Bhalerao, D., Jadhav, P. et al. (2020). Gut, oral and skin microbiome of Indian patrilineal families reveal perceptible association with age. Scientific Reports, 10(1), p.5685.

